# AN INTEGRATED APPROACH TO CARDIOVASCULAR DISEASE IN HOMELESS INDIVIDUALS: A QUALITATIVE STUDY

**DOI:** 10.1101/2022.11.20.22282560

**Authors:** Pippa Bark, Mel Ramasawmy, Andrew Hayward, Serena A Luchenski, Robert Aldridge, Stan Burridge, Amitava Banerjee

**Author notes:** Corresponding author: Prof Amitava Banerjee, Institute of Health Informatics, UCL, 222 Euston Road, London NW1 2DA, E.

## Abstract

**Background:** Homelessness is associated with an increased risk of cardiovascular disease (CVD), beyond impact of socioeconomic status. CVD is preventable and treatable, with barriers to both for homeless people. Those with lived experience of homelessness and health professionals with relevant expertise can help to understand and address these barriers.

**Objectives:** To understand and make recommendations to improve CVD care in homeless populations through those with lived experience and professional expertise.

**Method:** Four focus groups were conducted in March-July 2019. Three groups included people currently or previously homeless, each attended by a cardiologist (AB), a health services researcher (PB) and an “expert by experience” (SB) who coordinated participants. One group included multidisciplinary health and social care professionals in and around London to explore solutions.

**Participants:** The three homeless groups included 16 men and 9 women, aged 20- 60 years, of whom 24 were homeless and currently living in hostels and 1 rough sleeper. At least 14 discussed sleeping rough at some point.

**Results:** Participants were aware of CVD risks and relevance of healthy habits but identified barriers to prevention and health access, starting with disorientation affecting planning and self-care, lack of facilities for food, hygiene, and exercise, and experiences of discrimination.

**Conclusions:** CVD care for those experiencing homelessness should account for fundamental problems of the environment, be co-designed with service users and cover key principles: flexibility, public and staff education, integration of support, and advocacy for health service rights.

## BACKGROUND

Cardiovascular diseases (CVD) are the leading cause of disease burden globally[1]. Advances in prevention, diagnosis and treatment have reduced CVD mortality in high-income countries[2]. However, these improvements are unequally distributed, with persistent socioeconomic inequalities in CVD risk factors and morbidity[3], further exacerbated by social inclusion, including homelessness[4].

Research and policy addressing health needs of homeless populations have focused on infectious disease and crisis management[5,6]. While mental and behavioural disorders, and external factors are the most commonly recorded diagnoses at hospital admission, 95% of patients experiencing homelessness had a physical health need, in isolation, or in combination with mental illness and/or addiction[7]. CVD has a significant impact on health of homeless individuals, with higher prevalence, incidence and 1-year mortality risk, earlier onset, and a higher burden of risk factors than housed individuals [8].

Homelessness in the UK is rising, affecting over 270,000 people in 2021, of whom 2,700 were sleeping rough on any given night [9]. In the UK, NHS England has identified homelessness as a key area for action on health inequalities[10]. The case for CVD intervention is strong: managing acute disease and its consequences is expensive to the NHS[11]; and integrated approaches to CVD prevention and the potential for cost-saving have informed existing initiatives such as the NHS Health Checks[12]. In the population of individuals experiencing homelessness, this argument is further amplified - up to one third of deaths are from causes amenable to timely health care[13]; and due to ill-health, both causing, and being caused by, homelessness, this population is one of the highest cost groups for the NHS[14]. However, despite acknowledgement of the greater risk from CVD in this group, they have not been reached, due to factors such as poor GP registration rates, low health-seeking behaviour and high-risk younger individuals not being screened [15].

We aimed to identify barriers and facilitators to CVD care for homeless populations, drawing on those with lived experience of homelessness and health professionals.

## METHODS

### Participant recruitment

With support from local hostels and networks, a convenience sampling approach was applied to recruit individuals who were, or had been, homeless, in London and the South East of England. Homelessness was defined as including those in hostels and rough sleeping. Participants provided written consent.

### Data collection and analysis

Four focus groups were held between March 2019 and July 2019. Three focus groups were held in hostels with individuals who were, or had been, homeless: two with individuals currently residing in two London boroughs, and one with those outside London. Each group was attended by a cardiologist (AB), a health services researcher (PB) and expert by experience (SB) who coordinated the participants. A fourth focus group was held in London with health and social care staff working with homeless individuals. Interviews were digitally recorded and transcribed, and detailed notes were taken. Thematic analysis was carried out by PB and validated by AB. Themes were presented at a dissemination event 6 weeks after the focus groups and further validated.

### Ethics and PPIE

The study was fully approved by London – Camberwell St Giles Research Ethics Committee (18/LO/2153) on 21/12/2018. PPI representatives, co-ordinated by SB, contributed to the development of the NIHR Programme Development Grant (RP-DG-0117-10003), and design of the study including participant-facing materials. A dissemination event held in August 2019, was used to share and validate qualitative analysis and recommendations.

## RESULTS

Participants in the three groups with homeless individuals included 16 men and 9 women, of whom 24 were formally homeless and currently living in hostels and 1 was rough sleeping. Age was 20 to 60 years: 20 – 29 (n=5); 30 – 39 (n=7); 40 – 49 (n=10); and >50 (n=3) years. Participants in the professional group included 8 multidisciplinary staff from primary and secondary health and social care services in and around London, including cardiology, social work, pharmacists and homeless co-ordinators.

### Awareness of CVD within the homeless population

The groups presented a range of knowledge around causes of CVD. They had experience of parents and others having heart-related conditions, dying from heart attacks or following strokes, or unexpected deaths in those with no previous known history, and discussed hereditary factors. Whilst most did not volunteer having a heart condition initially, the typical background and lifestyle of rough sleeping, drug usage and smoking raised concerns of the long-term cardiac impact. Smoking was common, and not all had linked this to CVD. Having a cardiologist present provided an information source for a range of questions around personal health issues. The discussions revolved around concerns with current issues such as breathlessness, high blood pressure, palpitations and arrhythmia, and understanding helpful actions.

Whilst knowledge around diet, exercise, stress and smoking was high, participants discussed a number of direct and indirect barriers to health. The following themes explore the key concerns that relate to cardiovascular risk factors and health promoting behaviours: available facilities promote unhealthy eating habits; security and hygiene act as barriers to exercise; barriers to managing health conditions; the impact of stress and mental health, and coping mechanisms; and experience of discrimination in health services.

### Available food types and lack of access to food preparation space promote unhealthy eating habits

Although a healthy diet and fitness were understood as essential for better cardiac health, conditions of living and food donations made this much harder to achieve. Food was a priority, and consisted of “*death by sandwiches*”, cakes and “*party food*”. People reported putting on 2 or 3 stone whilst being homeless, with companies and members of the public routinely offering carbohydrates and sugar. One man summarised his own health: “*When I wasn’t homeless my weight say was between 71 kilos and 78. After my time on the streets and eating whatever I can I ballooned up to 120 kilos… it’s a struggle to get my weight back down again. I checked my heart rate on a monitor and my resting heart rate went up to 80-90 beats which is a lot higher than it should*.*”* A balance of healthier options was hampered by sugar cravings following substance abuse, tiredness or need for comfort, nowhere to cook food and the cost and availability of cheap junk food over healthier meals. Even in the hostels, people were given “*Fatty foods – we do get a lot of that where we are living. There are so many cakes. Cause it’s there we eat it. It tastes nice and you get overweight*.”

### Security and hygiene concerns act as significant barriers to exercise

Exercise was identified as a necessary factor in maintaining health, with some using walking and outdoor gyms as free options usually once in hostels. The more vulnerable talked about fear of walking away from where you were when sleeping rough in case you missed the chance of help and how the need to queue for a shower and to get food took up the majority of the day and personal energy.

“*When you’re homeless your daily struggle is to get a shower. So it means coming to places like this and waiting hours in the morning to get inside so your chances of exercise are diminished. You spend 3 or 4 hours in a queue waiting, then a couple of hours within a place literally stationary in a chair so you can do your bits and bobs and by the time you’ve come out and got a bit of food, its already 6 or 7pm*.”

Loss of previous activities such as boxing, yoga or gym meant not only reduced cardiac fitness but loss of companionship, normality, encouragement and peer network, where health related dialogue could have happened. Former exercise routines such as 5 –a-side football with friends were avoided because of the shame of arriving potentially smelling and/or not having appropriate kit. Support for exercise was welcomed, but only if in conjunction with hygiene facilities: *“But if you’re exercising, where can you get a wash or shower? It’s disgusting in the day centre*.”

### Barriers to managing existing health conditions

For those with a health condition, hygiene, managing appointments and taking regular medication was difficult. Medication could be stolen easily and it was difficult to store safely, keep dry, and to find private and hygienic places to take medication. Without a routine and with disturbed, fearful sleep, people become disorientated within days. They lost track of time and remembering appointments or collecting medication could be difficult. Mobile phones were seen as a solution but “*On the street not many have usable phones or they don’t have credit or they get stolen*.” A few participants had been given a £5 disposable health phone that helped.

### Impact of stress and mental health issues, and coping mechanisms are significant contributors to CVD

Stress and mental health issues contributing to CVD were a key discussion in all the groups. The conditions of insecure living, desperation and depression creating stress, stress increasing high blood pressure, and smoking, created the perfect storm. Someone commented that 75% of the homeless suffer mental health issues which are compounded by living on the street.

The majority smoked to alleviate stress and boredom and whilst giving up was discussed, the social norms in the hostels and perception of smoking as a stress reliever, meant that this was not seen as likely for most. “*I’ve quit before but take it up again because of stress. After giving up drugs and drugs, I smoke more. It’s a way of dealing with stress*.” Some pointed out that although there were public health initiatives, at this point in your life you could not action them without support. *“There are a lot of activities going down but some people if they’ve got that far down, they’re not able to cope on their own, and they need a big support system until they can have like a normal life again … Even with the benefits people are still going to smoke*.*”* Those who did not smoke tended to have an explicit reason for being different and whilst individuals talked about reducing, there was no obvious group norm to give up. Some commented that they had not realised that smoking damaged the heart, although they knew it was bad for health and presumably saw it as a lung issue in isolation.

We did not explicitly ask about alcohol or drug usage, however alcohol problems, marijuana, crack and cocaine came up when talking about the need to relieve stress or when asking about the effects of usage on the heart. Alternatives to stress relief were suggested by some individuals, as meditation and mindfulness, yoga, exercise and being outside. For others, life was still too chaotic to consider softer alternatives.

The timing of interventions for CVD prevention and management was also considered crucial. When sleeping rough, people talked about becoming disorientated within days or weeks, with lack of sleep, feeling unsafe and cold all contributing. “*The mental state of mind. You’re finding yourself homeless and in a different situation. Do you really think you’re going to be healthy? You want to drop dead. The last thing is health. They’re going to do drugs to forget what they do or get drunk and forget what’s happening to them*.” Although not all relied on substances, the sense of hopelessness was shared. Long term health issues were irrelevant at this stage and, with the sense of disorientation, “*responsibility and routine are not part of your life*.”

### Experiences of discrimination affect trust and belief in health services

Most of the participants (n=22) had a GP, but found attending on their own difficult. “Now I have a key worker, but I made every excuse not to go until I trusted her and slowly improved. Then I found her sitting in the GP holding my hand..” Moving address, sometimes frequently, also meant that the patient and the notes were separate “I had to wait 4 months. My records were in London and I was somewhere else. I could do day to day issues, but they didn’t know my history.”

Participants did not all realise you could change doctor when dissatisfied and many felt it would help to have a doctor in the day centre and more drop-in clinics: “*You need somewhere where you can find a doctor. You’re not going to keep an appointment*”. People liked having doctors who were specific to people dealing with people who were homeless. This was reflected in the discussions with the health and care professionals who noted that other professionals incorrectly perceived that standard health initiatives would not work in this group.

There was a strong sense that professionals treated the homeless differently. “Their attitude is different, their customer service is different. Everything is different. It’s as if they’re talking to an animal.” They described being left until last, discriminated against and treated like “dirt”. People perceived themselves as less likely to be heard: “If you’re in a shelter, you’re not recognised for the seriousness of what you’re saying. It took me ages to get heard. To get compassion.”

Being sent away from hospital because of actual or perceived drug abuse was common. “*In A&E, you get fobbed off because you’re homeless*”. Some individuals described making an effort to “*not to look homeless*” in order to be treated better. “*I’ve always been able to keep up my appearance so I don’t look too homeless… When I go to hospital, I need to make them like me*.”

People described getting a referral as ‘impossible’, with no reply to phone calls, and lengthy waits: “*I was at St Mary’s for an ECG for 2 hours and now I have to wait 9 weeks to see a GP*.” People talked about delays, some of which may have been specific to the homeless group. “*If you get in with these services, they are really good. If they believe you and that you are genuine. But you can spend months on the waiting list. There’s other people in more serious situations that need it*..” There had been several workarounds with people going to a dentist or optician as a way of getting a referral. One person talked about how the new duty to refer (a duty within the Homelessness Reduction Act 2017 for public authorities to refer those at risk or experiencing homelessness to local authority teams)[16] was going to make a difference, but it was taking time and most had not heard of it.

### Recommendations on improving cardiovascular disease care

CVD care has to be seen within the context of living conditions. Without shelter or support, many initiatives have no chance of being effective. The bottom line is housing. “*Do you think someone who has got a heart condition should be on the street? Someone who has congenital heart failure should be in a hospital or a house*.” Despite the legal obligation to help get people off the streets, people reported being on the street for months. It was once people were known and had deteriorated-“*That’s when they come looking for you and start doing things for you*.” Even following this, the groups reflected hostel living as the beginning, not the end of the journey. “*People when they come here, they don’t really care about their health. They need a push…*”

Focussing on CVD care, key themes for improving services included:

- **Flexibility of access to medical interventions** including: A mobile service that drops off prescriptions; safe medication storage; peer support to go to appointments; better referral avenues; and having doctors present at shelters and events for the homeless. It was identified that currently there were no health professionals there talking about heart health. A Find and Treat van linked to the current TB service was suggested as a potential avenue, but healthcare professionals raised concerns that this might also lead to the danger of overwhelming drop-ins with tests.
- **Supporting healthy behaviour through improved access** to hygiene facilities, exercise groups, and alternatives to stress reduction
- **Educating the public** for example, getting nutritional informational to people on streets and public wanting to help.
- **Informing those experiencing homelessness about their health service rights** such as: eligibility for annual GP check-up for the over 40s; and telling people about duty of care and how to get access or change GPs.
- **Increasing trust with the health system** through non-judgmental clinical staff, who are specialised at working with those experiencing homelessness
- Integrated substance abuse support

## DISCUSSION

This study provides insight into how those experiencing homelessness perceive and manage CVD. The participants expressed some awareness of the risk factors for CVD, and concerns for the impact of homelessness, such as rough sleeping, drug usage, smoking and mental illness, and stress, on their health. They identified actions that could be taken to manage this risk, but had limited control over these or facilitating factors. For example, although well-meaning, food provided was often unhealthy or sugary; and a lack of hygiene facilities, as well as insecurity of space and resources, contributed to difficulties in taking regular exercise. Negative experiences within the healthcare system further contributed to difficulties in managing existing or identifying new health problems. While recognising that immediate action required was access to safe shelter and support, participants had several recommendations on how CVD interventions could be made more approachable for this group. This included: flexibility of access to interventions (including safe medication storage, and accessible follow-up care); educating the public; advocating for health service rights; specialised and non-judgmental health and care staff; integrated substance abuse support; and supporting healthy behaviour through access to hygiene facilities, exercise groups, and alternatives to stress reduction.

### What is already known

The perception of ‘health’ in those experiencing homelessness has been characterised as a complex, and reaching beyond disease to encompass control over their life [17]. The need to balance competing priorities, such as access to food, shelter, and hygiene facilities, has been identified as barriers to health care access [18–21]. Additionally, stability of housing, and managing mental health issues or addiction to drugs or alcohol, are seen as pre-conditions to concentrating on a achieving a healthier lifestyle [17].

Previous studies with individuals experiencing homelessness have identified a good awareness of healthy behaviours and experience of similar barriers including: the type and availability of food and preparation facilities; access to exercise; and limited opportunities for stress reduction [17,22]. Smoking cessation has been described as something to address after their other priorities were resolved [17] – with social acceptability and pressure, behind preferences to reduce, rather than stop, smoking [23]. We identified an additional need to increase awareness of the cardiovascular impacts of smoking.

While a number of studies have focussed on the link between access to hygiene facilities and disease risk in the homeless population [24,25], fewer have focussed on the potential psychological impacts [26]. Inability to access water, sanitation and hygiene facilities causes stress and acts as a barrier not only to exercise [22], but to other higher-level needs such as securing employment [27]; and can be viewed as an important aspect of health itself in those experiencing homelessness [17].

Stigma and shame arising from lack of access to hygiene has also been described as a key feature in reluctance to access health services [17,28]; with experiences being described as shaped by ‘discrimination, disrespect and disempowerment’ [29]. Additional barriers to access have been characterised as: scattered organisations; difficult requirements to access care; care not adapted to their needs and not multidisciplinary; and a lack of continuity and planning [18,21,29]. Approaches that have been described as positive included the NHS Homeless Healthcare Team, which provides primary care services to the homeless, due to accessibility (co-location with other services, little waiting time, reminders about appointments and medication) [17].

Previous recommendations on improving access to care echo those put forward by the participants in our study. This includes building a trusting relationship, treating people with humanity and dignity; collaboration between professionals working in the care and support of those experiencing homelessness; flexibility within health services to meet user needs; training and support for professionals working with homeless people; and improved access to self-care facilities [18,29,30]. The need for practical solutions to improve medicine adherence, such as secure storage, has also been raised elsewhere [31]. Previous studies also support programmes that work directly with those experiencing homelessness to empower them with knowledge about their health and health service options, including clinic outreach [32], educational workshops and participatory research [33–35]; and health advocates [19].

### Strengths and limitations

This study provides additional insight into the prevention and management of CVD, a common cause of morbidity and mortality in those experiencing homelessness. In particular, it adds to the body of evidence calling for access to safe hygiene facilities as a key intervention to support health promotion and access to services. By including those experiencing homelessness and health and care professionals with expertise in supporting those experiencing homeless, our recommendations to improve CVD care are relevant and practicable in the context of local services.

By conducting research in London (and the South East), the study may not reflect the differences in experiences of homelessness across the UK, for example in rural areas, where it is rapidly rising [36]. Similar research is required for CVD and chronic diseases in homeless services in other countries.

## CONCLUSIONS

Cardiovascular disease care in those experiencing homelessness has to be seen holistically. Without shelter or support, many initiatives have no chance of being effective. The recommendations put forward by our participants include: flexibility of access to interventions; educating the public; advocating for health service rights; specialised and non-judgmental health and care staff; integrated substance abuse support; and supporting healthy behaviour through access to hygiene facilities, exercise groups, and alternatives to stress reduction. While services need to be co-designed with service users, and tailored to local conditions – for example, supporting existing services to integrate interventions and support for other diseases and health needs – consideration of these recommendations can promote service access and improve cardiovascular health in this population.

## Data Availability

Due to the nature of the research, supporting data is not available.

## Contributors

The research question was developed by AB. The study design and conduct were by AB, PB and SB. PB and MR wrote the original draft, with review and edits from AB. Additional review was carried out by SB, AH, SL and RA.

## Acknowledgements

We would like to thank the 25 men and women who gave us their time to share their experiences of rough sleeping and homelessness, which provided an invaluable insight into their specific health needs.

